# Emergence of Low-density Inflammatory Neutrophils Correlates with Hypercoagulable State and Disease Severity in COVID-19 Patients

**DOI:** 10.1101/2020.05.22.20106724

**Authors:** Samantha M. Morrissey, Anne E. Geller, Xiaoling Hu, David Tieri, Elizabeth A. Cooke, Chuanlin Ding, Matthew Woeste, Huang-ge Zhang, Rodrigo Cavallazzi, Sean P. Clifford, James Chen, Lu Cai, Maiying Kong, Corey T. Watson, Jiapeng Huang, Jun Yan

**Affiliations:** Department of Microbiology and Immunology, University of Louisville, KY, U.S.A.; Division of Immunotherapy, The Hiram C. Polk, Jr., MD Department of Surgery, Immuno-Oncology Program, James Graham Brown Cancer Center, University of Louisville, Louisville, KY, U.S.A.; Department of Biochemistry and Molecular Genetics, University of Louisville, KY, U.S.A.; Department of Anesthesiology and Perioperative Medicine, University of Louisville Hospital, KY, U.S.A.; Division of Pulmonary, Critical Care and Sleep Disorders, Department of Medicine, University of Louisville, Louisville, KY, USA; Pediatric Research Institute, Department of Pediatrics, University of Louisville, KY, U.S.A.; Department of Bioinformatics and Biostatistics, University of Louisville, KY, U.S.A.

## Abstract

Severe acute respiratory syndrome coronavirus 2 (SARS-CoV-2) is a novel viral pathogen that causes a clinical disease called coronavirus disease 2019 (COVID-19). Approximately 20% of infected patients experience a severe manifestation of the disease, causing bilateral pneumonia and acute respiratory distress syndrome. Severe COVID-19 patients also have a pronounced coagulopathy with approximately 30% of patients experiencing thromboembolic complications. However, the etiology driving the coagulopathy remains unknown. Here, we explore whether the prominent neutrophilia seen in severe COVID-19 patients contributes to inflammation-associated coagulation. We found in severe patients the emergence of a CD16^Int^CD44^low^CD11b^Int^ low-density inflammatory band (LDIB) neutrophil population that trends over time with changes in disease status. These cells demonstrated spontaneous neutrophil extracellular trap (NET) formation, phagocytic capacity, enhanced cytokine production, and associated clinically with D-dimer and systemic IL-6 and TNF-α levels, particularly for CD40^+^ LDIBs. We conclude that the LDIB subset contributes to COVID-19-associated coagulopathy (CAC) and could be used as an adjunct clinical marker to monitor disease status and progression. Identifying patients who are trending towards LDIB crisis and implementing early, appropriate treatment could improve all-cause mortality rates for severe COVID-19 patients.

**One Sentence Summary:** In this study, we discover that low-density neutrophils significantly contribute to COVID-19-associated coagulopathy and inflammation

## Introduction

December 2019 saw the emergence of a novel viral pathogen, severe acute respiratory syndrome coronavirus 2 (SARS-CoV-2). To date, there have been over 4 million cases worldwide with upwards of 300,000 reported deaths(*1*). SARS-CoV-2 is considered a lower respiratory tract pathogen that gains access to the body by binding to the angiotensin-converting enzyme 2 (ACE-2) on the surface of alveolar epithelial type II cells(*2*). The virus causes a clinical disease called coronavirus disease 2019 (COVID-19)(*3*). While the majority of persons infected with COVID-19 experience mild to moderate symptoms of pharyngitis, rhinorrhea, and low-grade pyrexia, approximately 20% of patients experience a severe influenza-like manifestation of the disease(*3*). Clinically, these patients present with bilateral pneumonia progressing to acute respiratory distress syndrome (ARDS) with a marked decreased in pulmonary function requiring mechanical ventilation(*3-5*). The fluid accumulation in the lungs that is pathognomonic for ARDS results from a combination of virally induced lung injury as well as the rapid influx of immune cells to fight the infection(*6*). These recruited inflammatory mediators are often in a hyper-activated state causing a phenomenon known as cytokine storm(*7*). There have been a variety of cytokines associated with cytokine storm including interleukin-6 (IL-6), interleukin-1β (IL-1B), and tumor necrosis factor-α (TNFα)(*8*). If the high levels of cytokines go unresolved, patients are at an increased risk of vascular hyperpermeability, multi-organ failure, and death(*9*). Levels of all three cytokines have been found to be elevated in the peripheral blood of COVID-19 patients(*3, 10, 11*).

Severe COVID-19 patients have a distinct immunological phenotype characterized by lymphopenia and neutrophilia. Patients with an increased neutrophil to lymphocyte ratio (NLR) have reported worse clinical outcomes(*10*). Lung specimens at autopsy showed a marked infiltration of neutrophils into the lung tissue(*12, 13*). Neutrophils are thought to be recruited to the lungs to aid in the clearance of the viral pathogens through phagocytosis, secretion of reactive oxygen species, and cytotoxic granule release(*14*). However, prolonged activation of these neutrophils has been linked to adverse outcomes in patients with influenza. Specifically, neutrophil populations in patients with severe H1N1 influenza infection showed increased extracellular net formation, neutrophil mediated alveolar damage, and delayed apoptosis(*15, 16*). These factors predominately contributed to mortality in animal models of the disease.

In addition to significant pulmonary complications, severe COVID-19 patients also have a notable coagulopathy(*17, 18*). Multiple studies report COVID-19 patients experiencing thromboembolic events including myocardial infarction, pulmonary embolism, cerebrovascular accident, and deep vein thromboses(*19, 20*). The majority of patients with severe disease have increased D-dimers, platelet abnormalities, and decreased prothrombin time (PT) or partial thromboplastin time (PTT) over the course of their hospitalization(*21*). Given the prevalence of thromboembolic complications in severe COVID-19 patients, the standard of care for intubated patients now includes full anticoagulation therapy(*9, 17*). However, the etiology of the coagulopathy has yet to be clearly elucidated. In this study, we investigate the hypothesis that the excessive neutrophilia seen in severe COVID-19 patients directly contributes to COVID-19-associated coagulopathy (CAC). We found that the most severe patients, requiring mechanical ventilation, demonstrated a marked increase in the overall CD66b^+^ neutrophil percentage within the peripheral blood compartment as compared to moderate patients. Within the severe COVID-19 patient cohort, we also saw the emergence of a significant population of CD16^Int^CD44^Low^CD11b^Int^ low-density neutrophils, which we refer to as low-density inflammatory band cells (LDIBs). The increases in this population trended with disease severity and mortality while decreases were associated with extubation and discharge. Additionally, the LDIB population percentage trended with D-dimer levels across all COVID-19 patients. Functional analysis of these cells revealed their phagocytic activity, spontaneous formation of neutrophil extracellular traps (NETs), and enhanced secretion of IL-6 and TNF-α. Plasma levels of IL-6 in all COVID-19 patients positively correlated with the LDIB population while TNF-α showed a trending correlation. Taken together, these findings suggest that LDIBs significantly contribute to CAC. This study will hopefully help clinicians to better delineate which patients are at the highest risk for developing thromboembolic complications and determine when to treat with appropriate immunomodulatory agents.

## Results

### Neutrophil profiling in hospitalized COVID-19 Patients

For our study, we enrolled a total of 13 patients that had tested positive for SARS-CoV-2 by nasopharyngeal swab. Seven patients were initially enrolled in the severe category as defined by necessity of mechanical ventilation within the intensive care unit (ICU) and six were initially enrolled in the moderate group, as patients that had been admitted to the hospital but were not on a mechanical ventilator. The patient demographics was summarized in Table 1. The average age of COVID-19 patients was 66.8 with a male to female ratio of 8:5. Of note, 5/7 severe patients (71.4%) and 3/6 moderate patients (50%) experienced a thromboembolic complication either as a presenting illness or during the course of their hospitalization. Peripheral blood samples were drawn daily from either a venous or arterial line for severe patients whereas moderate patients had samples drawn from a venous line approximately every two to three days.

**Table 1.** Study participant demographics.

|  | **Total** | **Healthy** | **COVID-19** |
| --- | --- | --- | --- |
| Participants | 19 | 6 | 13 |
| Age, mean, years | 61.26 (28-95) | 50 (28-68) | 66.8 (28-95) |
| Male: Female | 12:7 | 4:2 | 8:5 |
| Race |  | 5 Caucasian<br>1 Asian | 8 Caucasian<br>5 Black/ African American |
| Mean Comorbidities, St<br>Deviation |  | .3 ± .47 | 4 ± 2.0 |
| Patients experiencing<br>thromboembolic<br>complications during<br>hospital stay |  |  | 8 (61.5%) |
| Patients receiving<br>Hydroxychloroquine +<br>Azythromycin |  |  | 6 (46%) |
| Patients receiving<br>convalescent plasma |  |  | 4 (30.7%) |
| Mortality |  |  | 4 (30.7%) |

We began our study by comparing the CD45^+^ lineage clusters between healthy donors, moderate, and severe COVID-19 patients (Figure S1a). Cell lineage cluster analysis demonstrated that CD66b^+^CD16^+^ neutrophils (cluster 1, Figure S1b) were the most prominent population in COVID-19 patients which agrees with previous reports indicating a dominant neutrophilia in these patients(*10*). We confirmed our results with data pooled from patient serial whole blood complete blood count (CBC) reports. This data demonstrated that severe patients had approximately a 10% increase in neutrophil percentage in their peripheral blood as compared to moderate patients, and a 30% increase over healthy donors (Figure 1a). Conversely, the overall lymphocyte percentage in these patients was decreased as compared to the moderate cohort and healthy donors. viSNE analysis of the overall CD3^+^ T cells and CD4^+^ and CD8^+^ T cell subsets showed decreasing population size in patients with moderate and severe COVID-19 as compared to healthy donors (Figure S2a). Taken together, this data ultimately characterizes an increased NLR within our severe cohort (Figure 1a) that agrees with previously published reports(*22*).

**Fig. 1.**
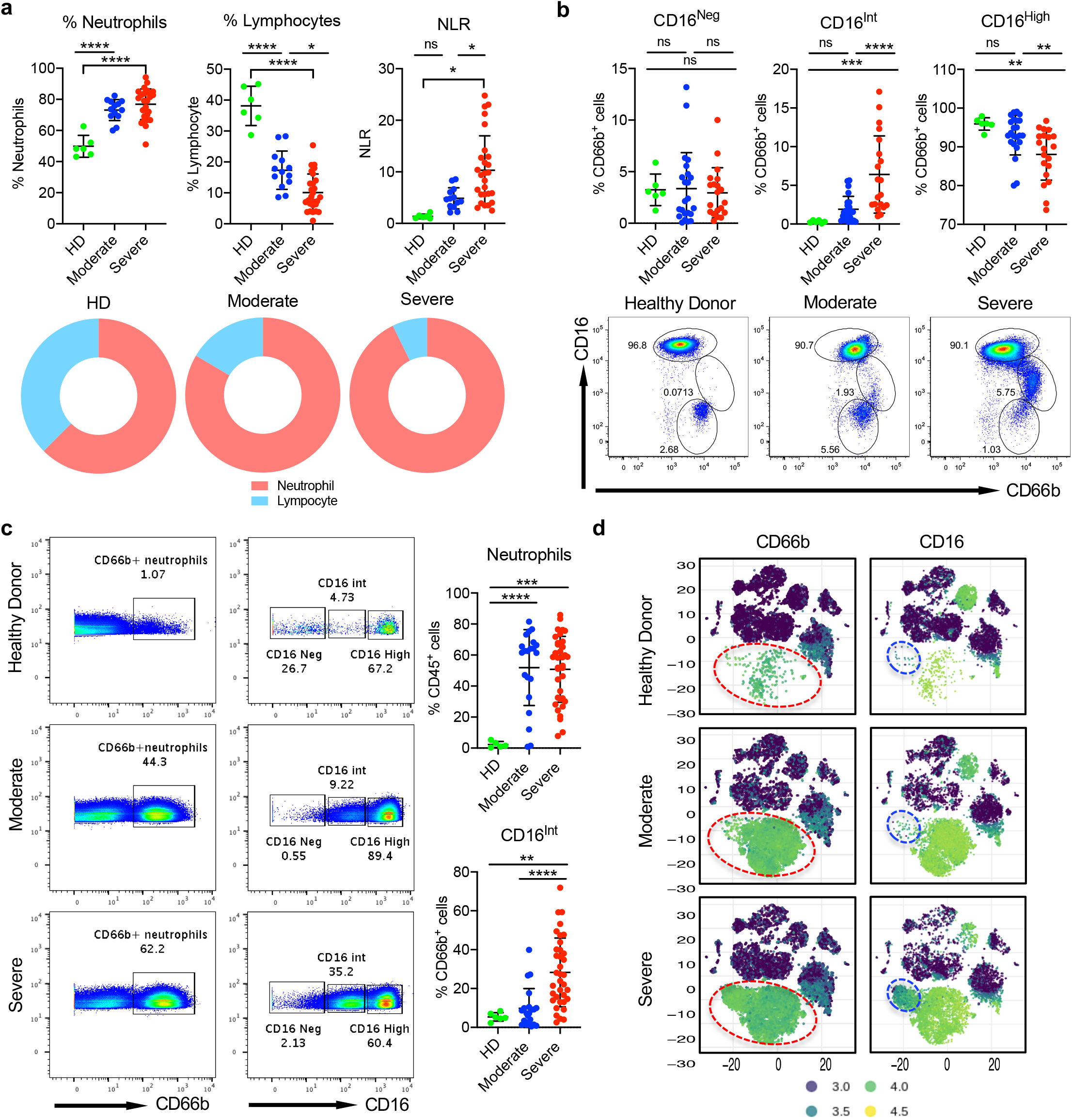
The identification of a CD16 intermediate low-density neutrophil population in COVID-19 patients. **(A)** Neutrophil and lymphocyte percentages and the neutrophil to lymphocyte ratio in whole blood as measured by a clinical complete blood count (CBC) in HDs and patients with moderate and severe COVID-19 infection. Data are pooled from serial blood samples collected from 5 HDs, and serially from 6 moderate patients and 7 severe patients starting from the day of enrollment. Each draw from each patient represents one data point and is related to the condition of the patient (moderate or severe) on that day. HD (n=6), Moderate timepoints (n=13), Severe timepoints (n=27). Pie charts depict representative data of the neutrophil to lymphocyte ratio (NLR) in HDs, severe and moderate patients. **(B)** The percent of CD16 negative (CD16^Neg)^, CD16 intermediate (CD16^Int^), and CD16 high (CD16^High^) neutrophils from whole blood samples among HD (n=5), moderate (n=22), and severe (n=30) serially drawn COVID-19 samples. Samples are gated the CD45^+^CD66b^+^ population and show an increased CD16^Int^ population in moderate and severe COVID-19 patients. Summarized data and representative dot plots are shown. **(C)** Representative dot plots (left) and summarized data (right) showing the overall percent of CD66b^+^ neutrophils (gated on viable, CD45^+^) as well as CD16^Neg^, CD16^Int^, and CD16^High^ subsets as found in Ficoll isolated PBMCs analyzed using CyTOF mass cytometry in healthy donors (n=5), moderate samples (n=21) and severe samples (n=36). **(D)** Representative viSNE cluster plots generated using CyTOF work flow show the CD45^+^ PBMC populations in HDs, and patients with moderate and severe COVID-19. Plots highlight an increased intensity of the CD66b^+^ neutrophil population (left) and CD16^+^ populations (right) in HDs versus moderate and severe COVID-19 patients. Red circles indicate the location of the neutrophil population while the blue circle indicates the CD16^Int^ population. In all summarized data, the mean with standard deviation is represented. p values were determined using a linear mixed effect model. ns= p ≥.05, * p< 0.05, ** p<0.01, **** p < 0.0001.

Further investigation into the neutrophil pool revealed three distinct subpopulations within whole blood samples that clustered by CD16^Neg^, CD16^Int^, and CD16^High^ expression. Severe COVID-19 patients showed a marked increase in the CD16^Int^ subset, which was significantly lower in the moderate cohort, and virtually absent in the healthy donors (Figure 1b). CD16^Int^ neutrophils classically have been reported to be low-density neutrophils or immature neutrophils(*23*). Clinically, immature neutrophils are called band cells and are associated with a left shift on a complete blood count (CBC)(*24*). These neutrophils are often mononucleated and smaller than typical neutrophils. Therefore, due to the combination of their number and smaller mononucleated morphology, we were able to pull down a significant portion of these cells from the blood using a typical PBMC Ficoll isolation method. Previous reports also described this phenomenon in more severe cases of autoimmunity(*25*). Minimal neutrophils were isolated from healthy donors using this method indicating the unique characteristics of these LDIBs in COVID-19 patients. Isolating the LDIBs via Ficoll resulted in about ~6-fold enrichment of these cells over peripheral blood within each cohort (Figure 1c). Therefore, while the actual percentage was higher than in whole blood (Figure 1b), the ratio between the cohorts was similar thus allowing for valid comparisons. Cluster analysis of isolated PBMCs from a single blood draw in each donor indicated a predominate neutrophil population (circled in red) within the CD45^+^ compartment in the severe COVID-19 cohort as compared to moderate patients and healthy donors (Figure 1d, left panel). Additionally, in the severe patients, there was a subset of the neutrophil population that expressed intermediate CD16 (blue circle) which was diminished in both the moderate and healthy donors (Figure 1d, right panel). This adjacent CD16^Int^ cluster represented the LDIB population seen in severe COVID-19 patients (Figure 1c).

Interestingly, tracking the CD16^Int^ LDIB population over the course of each patients’ individual hospital stay revealed an important association between clinical outcomes and the percentage of CD16^Int^ neutrophils (Figure S2b). Specifically, in patients 3, 4, and 5, the percentage of CD16^Int^ cells trended with improvements in disease status. As their CD16^Int^ percentage began to decline, these patients were extubated and switched from the severe to moderate group. Conversely, in patients 1, 8, 12, patient mortality was directly associated with an increasing CD16^Int^ percentage as compared to their baseline at enrollment or the CD16^Int^ neutrophil percentage stayed constantly at a high level (patient 1). Lastly, patients 6, 7, and 9 in the moderate group consistently had a minimal CD16^Int^ population for the duration of the hospitalization prior to their discharge. Collectively, these findings suggest that the most severe COVID-19 patients experienced an emergence of LDIB population that trends with both improvements and declines in patient status.

### Phenotypic characterization of CD16^Int^ LDIB cells

Maturation of neutrophils from hematopoietic stem cells is identified by stages with distinct morphological characteristics(*26*). We performed Wright-Giemsa staining to determine if the three CD16 populations of neutrophils were actually neutrophils in the later three stages of development: myelocyte, metamyleocyte (band cell), and granulocyte (mature neutrophil). Figure 2a clearly showed that the CD16^Neg^ cells were basophilic myelocytes with an ovoid nucleus, the CD16^Int^ cells were band cells with the characteristic band shaped nucleus, and the CD16^High^ cells were segmented, mature neutrophils. However, it is relevant to note that the mature CD16^High^ neutrophils are bi-lobed rather than hyper-segmented and closely resemble pseudo-Pelger-Huet cells. Pseudo-Pelger Huet cells have been described in other severe infections like influenza A, tuberculosis, and human immunodeficiency virus (HIV)(*27*). It has been suggested that these cells develop as a result of excessive exposure to inflammatory factors like TNF-α and IFN-γ(*28*).

**Fig. 2.**
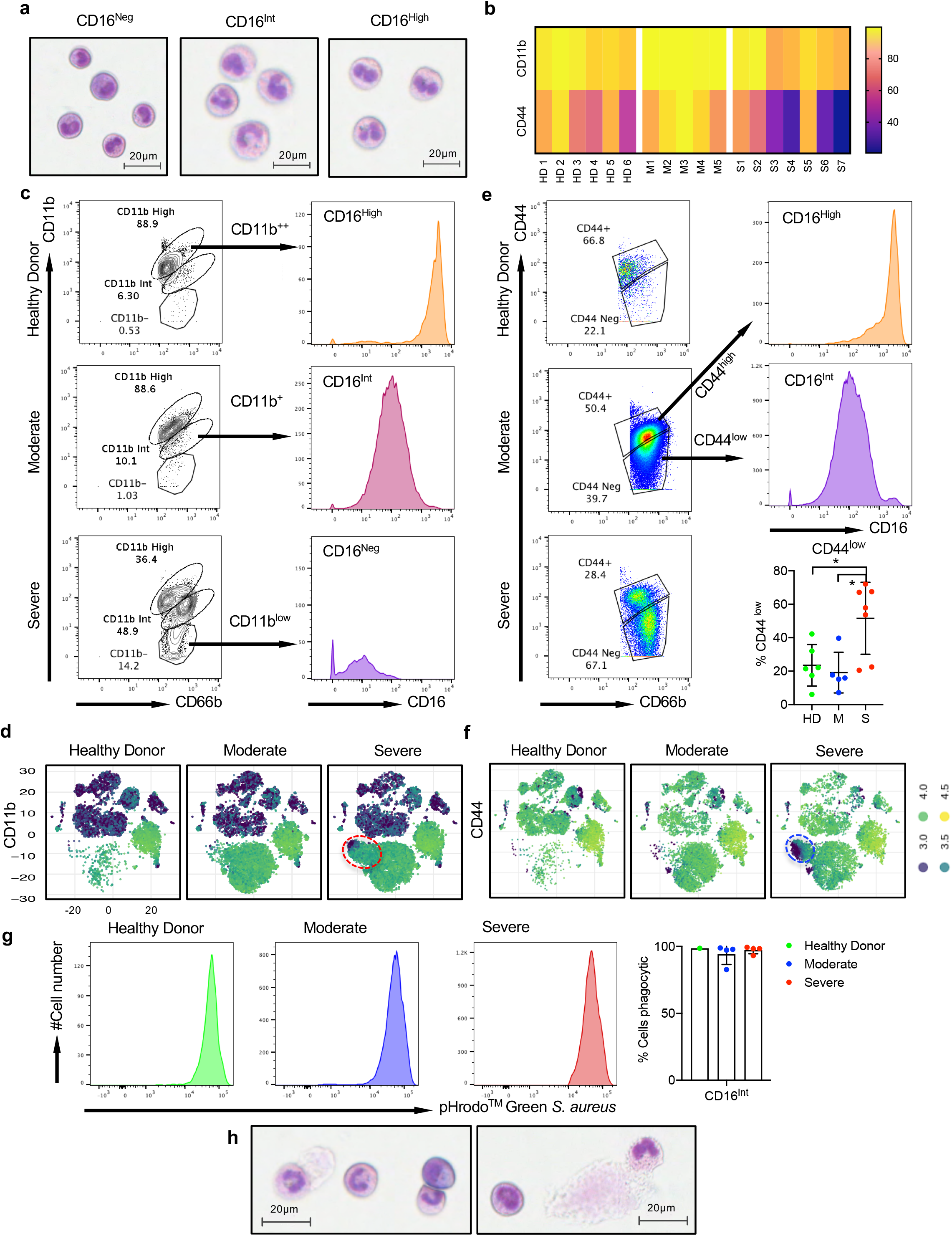
Phenotypic characteristics of CD16^Neg^, CD16^Int^, and CD16^High^ neutrophil populations. **(A)** Wright Giemsa staining of CD66b^+^ CD16^Neg^ (left), CD16^Int^ (middle), and CD16^High^ (right) populations that were enriched using Fluorescence Activated Cell Sorting (FACS) show different stages of neutrophil maturation. **(B)** The heatmap shows differential expression of CD11b (top) and CD44 (bottom) on CD66b^+^ neutrophils in Ficoll isolated PBMCs analyzed via mass cytometry. Here, 6 HDs, 5 moderate COVID-19 patients and 7 severe COVID-19 patient samples from the first day of study enrollment were used. **(C)** Using mass cytometry, CD11b expression on CD66b^+^ neutrophils segregated into three distinct populations: CD11b high (CD11b^++)^, CD11b intermediate (CD11b^+^) and CD11b low (CD11b^-^). CD11b^++^cells were found to be CD16^high^ (top), CD11b^+^ cells were found to have intermediate CD16 expression (middle) and CD11b” cells showed low CD16 expression (bottom). **(D)** viSNE cluster plots generated using CyTOF work flow highlight the expression of CD11b in the CD16^Int^ neutrophil population (indicated by the red circle). An increase in the CD11b^+^ population can be seen in moderate and severe COVID-19 patients as compared to HDs. **(E)** Using mass cytometry, CD44 expression on CD66b^+^ neutrophils segregated into two distinct populations: CD44 positive (CD44^high^) and CD44 negative (CD44^low^). The CD44^high^ population is shown to have high expression of CD16 as shown by the histogram, while the CD44^low^ population is shown to have intermediate expression of CD16. Summarized data includes the first sample acquired from each patient enrolled in the study, and shows that the percent of CD66b^+^ CD44^low^ neutrophils is significantly increased in severe patients (n=7) as compared to HDs (n=6) and moderate patients (n=5). Statistics were performed using a one-way ANOVA where * p<.05 **(F)** viSNE cluster plots represent the decreased expression of CD44 in the CD16^Int^ neutrophil compartment in severe COVID-19 patients as compared to moderate patients and HDs, as highlighted bythe blue circle. **(G)** The phagocytic capacity of neutrophils from whole blood in HDs, severe and moderate patients was assessed using a pHrodo™ Green E.Coli BioParticles™ phagocytosis assay. Representative histograms show the relative phagocytic capacity of CD16^Int^ populations in HD (left), moderate (middle), and severe patients (right), and summarized data indicates the percent of phagocytic cells in the CD16^Int^ population, HD (n=1), moderate (n=4) and severe (n=4). **(H)** Wright Giemsa staining of CD66b^+^ neutrophils showed spontaneous NET formation from CD16^Int^ LDIB neutrophils.

Next, we explored differential surface marker expression on the different CD16 subsets of neutrophils in COVID-19 patients. We first performed cluster analysis on the overall CD66b^+^ neutrophil population. As shown in Figure S3a, there was an increased prevalence of cluster 2 in the severe patient cohort as compared to moderate and healthy donors. Conversely, there was a slight decrease in density of cluster 1 in the severe group as compared to the other two. Utilizing the heatmap in Figure S3b revealed that cluster 1 expressed high levels of CD11b, CD44 and CD16. Conversely, cluster 2 showed decreased expression of CD44, CD16, and CD11b. Interestingly, tracking the neutrophil clusters in serial blood draws over 5 days from different types of patients revealed the dynamic nature of neutrophil pools in COVID-19 infection (Figure S4a, b). In the severe patient, over the time, the light blue population (cluster 4, black circle) increased while all the other clusters remained similar. For the moderate patient, the majority of clusters remained stable over time. The patient that was initially enrolled in the severe cohort but changed to moderate by day 5, had a profound decrease in cluster 5 (red circle) over time. Conversely, in the patient that transitioned from moderate to severe, the light blue (cluster 4) and purple clusters (cluster 5) increased over the time, which was consistent with the change in disease severity (Figure S4a).

Understanding that the profile of neutrophil clusters associates with disease status, we wanted to expand upon the findings from our analysis and determine a specific surface marker phenotype for three CD16 neutrophil clusters. To do this, we generated a heatmap from the CyTOF analysis profiling the CD66b^+^ population within the three cohorts (Figure 3a). Two markers from our cluster analysis, CD11b and CD44, stood out to be differentially expressed between the healthy donors and the two patient cohorts (Figure 2b). CD11b expression level was intermediate in the severe cohort while CD44 was the lowest in this patient population. Breaking CD11b expression down by CD16 subset, showed increasing expression of CD11b as the cells progress through the developmental stages, with the LDIBs having an intermediate expression profile (Figure 2c). Cluster analysis revealed that the representative LDIB cluster indeed showed decreased CD11b expression (red circle) as compared to the overall CD66b^+^ neutrophil cluster (Figure 2d).

**Fig. 3.**
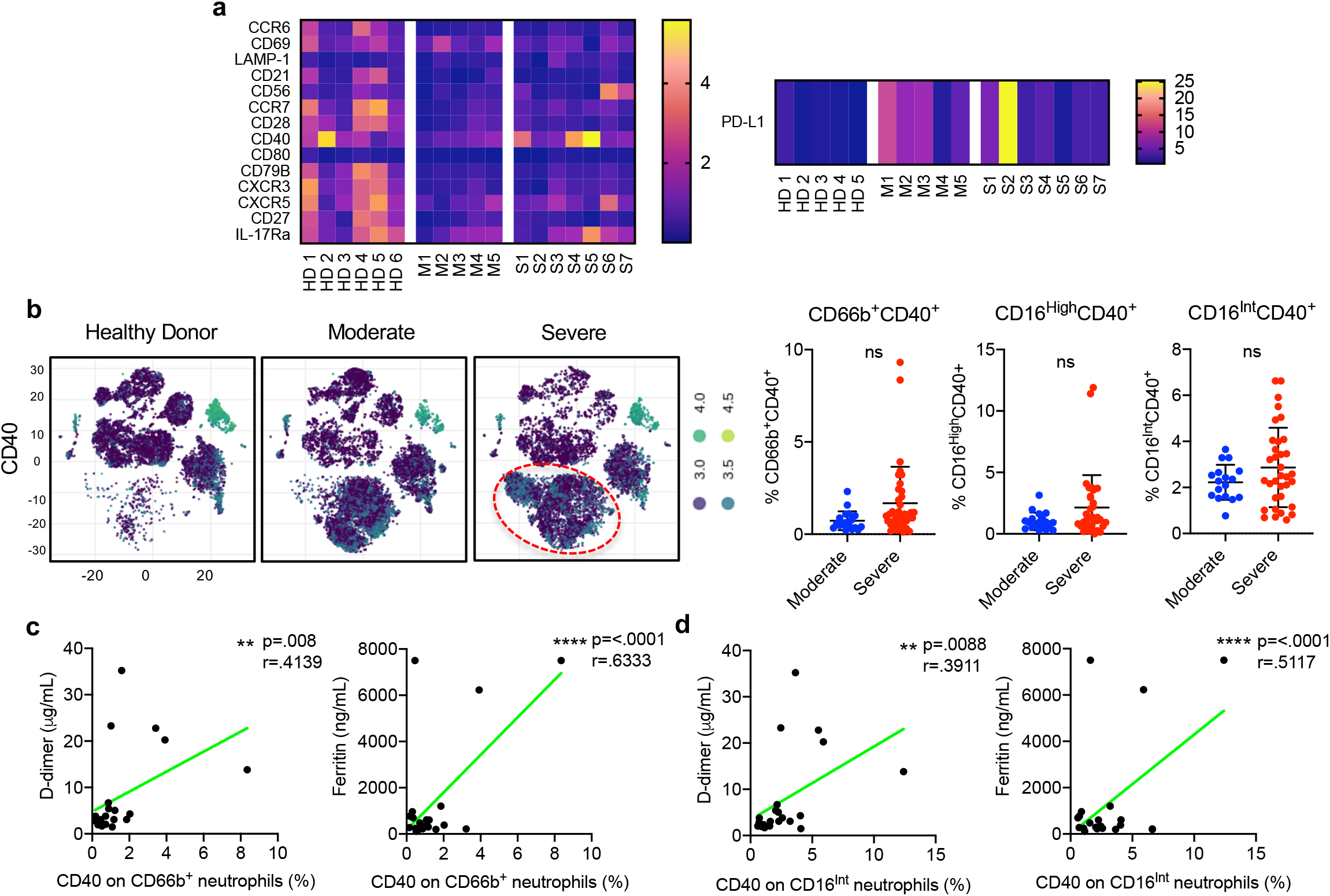
The expression of CD40 on neutrophils and correlation with clinical measures of coagulation. **(A)** Heatmaps showing the overall expression of various surface markers on the CD66b^+^ neutrophil population in HDs (n=5), moderate (n=5) and severe (n=6) patients on their first day of study enrollment. **(B)** Representative viSNE plots showing increased CD40 expression on the overall CD66b^+^ neutrophil population (indicated by the red circle) healthy donors, moderate and severe COVID-19 patients (left). Summarized expression of CD40 on the overall neutrophil pool as well as on the CD16^High^ and CD16^Int^ neutrophil subsets in COVID-19 patients (right). Data were pooled from serial patient draws throughout the course of their hospital admission and grouped according to patient status. A linear mixed effect model was used to determine significance. **(C, D)** D-dimer (n=22) and ferritin (n=21) values from serial samples from the severe cohort only were correlated with the percent of CD40^+^CD66b^+^ total neutrophils **(C)** and the percent of CD40^+^CD16^Int^ neutrophils **(D)**. Marginal Pearson correlations were used to indicate statistical significance in all correlations, where ** p< 0.01, **** p<0.0001.

CD44 is an important surface marker that has been associated with neutrophilic lung inflammation in bacterial pneumonia. Decreased surface expression of CD44 resulted in increased accumulation of neutrophils in the lungs of *E.coli* infected mice(*29*). Therefore, given the known accumulation of neutrophils in the lungs of severe COVID-19 patients, it was not surprising that the CD16^Int^ cells had the lowest expression of CD44 indicating the highest potential for infiltration into the lung (Figure 2e). Cluster analysis further confirmed these findings (Figure 2f, blue circle). Since CD44^Low^ neutrophils are recruited to the lung to aid in clearance of bacterial pneumonia, we next investigated the phagocytic properties of the neutrophils from COVID-19 patients. Figure 2g showed that CD16^Int^ LDIB cells had a high uptake of pHrodo green *S.aureus* bioparticles suggesting a highly activated phenotype. One of the main ways that neutrophils eliminate pathogens is through NETs, the extravasation of DNA and protein to form a web like structure that can trap and kill extracellular pathogens. Increased NET formation from neutrophils in mouse models of bacterial sepsis increased platelet aggregation and coagulation(*30*). During analysis of the Wright-Giemsa stain for neutrophil morphologic characterization, we noticed that the LDIBs were spontaneously forming NETs more prominently than CD16^Neg^ or CD16^High^ (Figure 2h). Previous reports have also noted that low-density neutrophils readily form NETs causing endothelial vessel and organ damage in autoimmune phenotypes, which further confirms the pathogenic role of LDIBs in COVID-19(*25*).

Another neutrophil factor besides NETs that has been associated with driving platelet activation and thrombosis is CD40. Inhibition of the neutrophil-platelet CD40/CD40L axis with anti-CD40 antibody significantly diminished pulmonary edema, platelet activation and neutrophil recruitment to the lungs in a mouse model of transfusion related acute lung injury (TRALI)(*31*). Assaying for CD40 expression on the neutrophil subsets, we found the overall neutrophil population in severe patients had a trending increased expression of CD40 as assessed by cluster analysis and flow cytometry (Figure 3b, red circle) although not statistically significant. Strikingly, CD40 expression on the total neutrophils and CD16^Int^ LDIB population significantly positively correlated with severe COVID-19 patients’ D-dimer and ferritin levels (Figure 3c, d), suggesting a potential involvement of severe inflammation and thrombus formation.

### Clinical significance of LDIB neutrophils in CAC

Understanding the etiology of CAC is of paramount importance so that early adjustments in clinical management can be made to improve overall survival outcomes. Anti-coagulation therapy has been shown to increase the overall survival of both non-ventilator and ventilator dependent COVID-19 patients(*32*). However, anti-coagulation therapy comes with risk and is contraindicated in some patients(*19*). Therefore, it is necessary to delineate which patients are at the highest risk for thromboembolic complication and determine other potential strategies to mitigate inflammation induced coagulation in these patients.

Two of the main clinical markers used to monitor coagulation state are D-dimer and platelet count, where increased D-dimer levels and decreased platelet counts are associated with coagulation(*17*). Looking into our COVID-19 cohort, we found that severe patients had an elevated level of D-dimer compared to moderate patients (Figure 4a). The platelet levels were also increased in severe patients. Two other clinically important markers used to monitor systemic inflammation are ferritin and lactate dehydrogenase (LDH) (*33*). While ferritin was not different between the two groups, it was elevated in moderate and severe COVID-19 patients as compared to the normal range(*34*). Increased LDH levels as seen in the severe cohort were often associated with more severe lung damage and tissue injury(*33*).

**Fig. 4.**
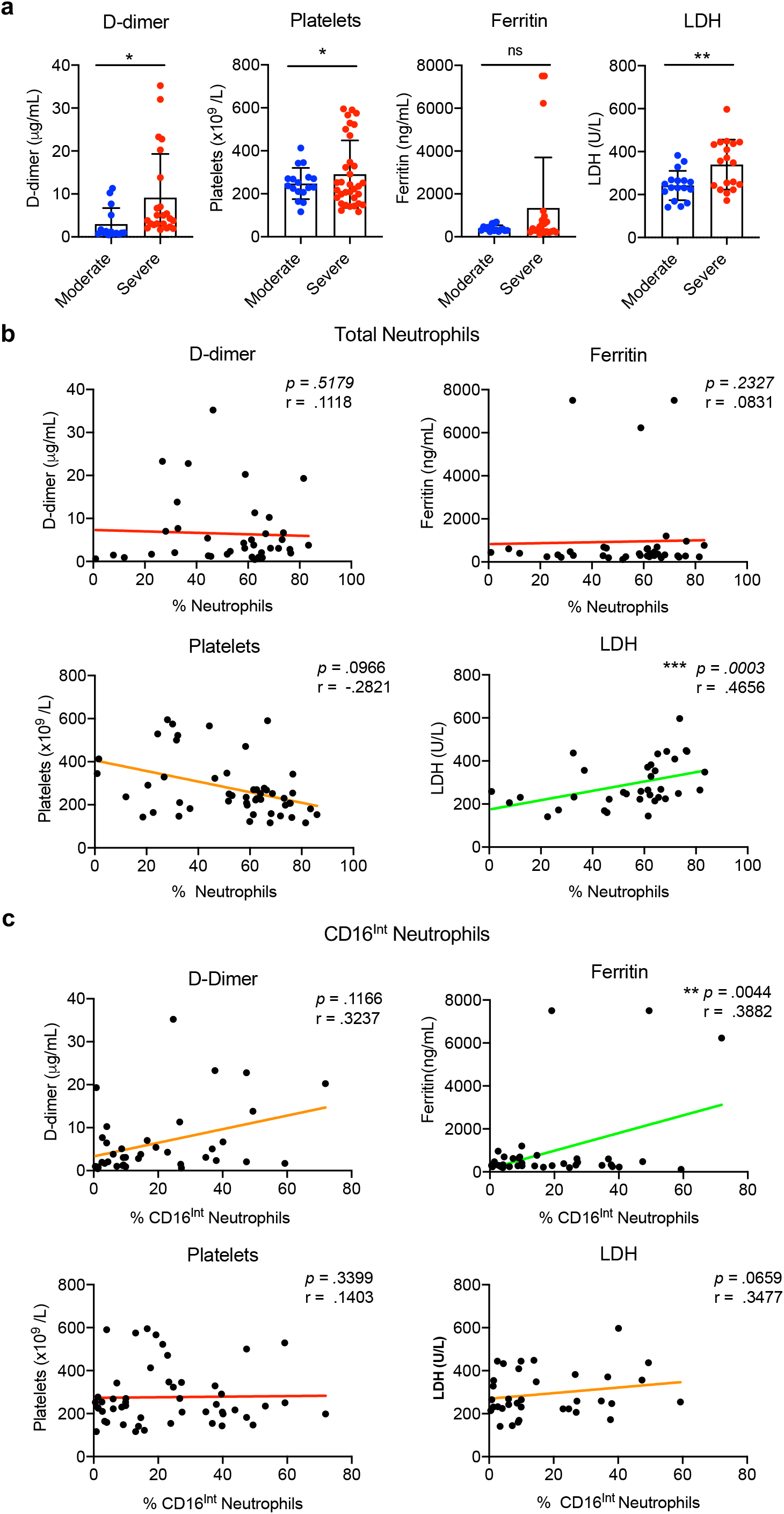
Correlation of clinical coagulation indicators with neutrophils and LDIBs. **(A)** For severe and moderate patients, the clinical values of D-dimer, Ferritin, Platelets and LDH were acquired from patient charts, and serial blood draws from patients were grouped based on patient status. These values were recorded approximately every other day during hospital admission and were pooled to generate summarized data. D-dimer samples: moderate (n=15), severe (n=23), Ferritin samples: moderate (n=16), severe (n=22), Platelet samples: moderate (n=17), severe (n=33), LDH samples: moderate (n=17), severe (n=18). A linear mixed effect model was used to determine significance. * p<.05 ** p<0.01 **(B)** The D-dimer (n=38), ferritin (n=38), platelet (n=50) and LDH (n=35) levels for all COVID-19 patient samples in Figure3a were correlated with the total neutrophil percentage in the Ficoll isolated PBMCs on the day of that charted measurement. **(C)** The D-dimer, ferritin, platelet and LDH values (n=same above) were correlated with the corresponding percent of CD16^Int^ neutrophils in the Ficoll isolated PBMCs found on the same day at the clinical reading. For all correlation data, a line of best fit is shown to visually examine correlation, with a green line representing a statistically significant correlation, a red line representing a non-significant correlation and an orange line representing a trending correlation that was not significant. Marginal Pearson correlations where used to indicate statistical significance in all correlations, where ns= p ≥.05, ** p<0.01, *** p<0.001.

Having a better understanding clinically of the relevant markers in our two patient cohorts, we first sought to determine if overall neutrophil percentage was a good diagnostic tool to determine high risk of thromboembolic event. Figure 4b showed that overall neutrophil percentage did not correlate with D-dimer or ferritin levels. However, overall neutrophil percentage did negatively trend with platelet counts and positively correlate with LDH levels suggesting some association with thrombosis and declining status. Conversely, the CD16^Int^ population significantly correlated with ferritin but not platelets or LDH (Figure 3c). For correlation with D-dimer, we clearly saw a trend between the LDIB population and D-dimer, although statistical significance was not reached (Figure 4c). Two issues related to this analysis were that D-dimer level was not measured frequently in our cohort of patients, particularly for moderate patients and all patients received anti-coagulation therapy (Table 2). However, despite this, trending serial analyses of individual patients’ LDIB populations with D-dimer demonstrated appreciable associations and a pronounced phenotype. Taking patient 12 as a representative severe patient, there was a clear correlation between their rising D-dimer levels and increasing LDIB population leading up to their death (Figure S5a). Alternatively, in patient 9 from the moderate group, both their D-dimer and LDIB percentage were only marginally elevated prior to discharge (Figure S5b). Therefore, taking the statistical and descriptive data together our finding suggests that the LDIB percentage rather than overall neutrophil percentage correlates better with coagulation status in COVID-19 patients.

### Contribution of LDIBs to cytokine-mediated coagulopathy in COVID-19 patients

It has been established that severe COVID-19 patients have elevated levels of pro-inflammatory cytokines resulting in cytokine storm(*3, 9*). Two of the main cytokines that have been found to be consistently elevated among the most severe COVID-19 patients are TNF-α and IL-6(*3, 35*). In cytokine storm, TNF-α causes vasodilation and increases vascular permeability to allow for immune infiltration, resulting in pulmonary edema(*36*). IL-6 induces a multitude of immunmodulatory functions including T cell and B cell activation, acute phase reactive protein production from the liver, and platelet hyper-activation (*37, 38*). Both IL-6 and TNF-α have been reported to promote coagulation through activation of the extrinsic coagulation cascade by inducing endothelial expression of tissue factor(*39*). Therefore, given the associations between IL-6 and TNF-α with cytokine storm and coagulation, we wanted to determine if LDIBs and/or overall neutrophils were contributing to the generation of these cytokines and whether they correlated with clinical markers of coagulation.

We first measured plasma concentrations of TNF-α and IL-6 in the serial blood samples of patients compared to healthy donors (Figure 5a). The overall plasma level of TNF-α was low but was elevated in the severe group compared to moderate and healthy donors. IL-6 showed significant increases above moderate patients. Correlating the plasma level of TNF-α with overall neutrophil percentage showed no significant association while IL-6 level was significantly correlated with overall neutrophil percentage (Figure 5b). Furthermore, the CD16^Int^ LDIB population showed a positive significant correlation with IL-6 levels across all patients and donors and TNF-α level showed a strong trend with LDIB frequency. These results further emphasize the particular pro-inflammatory characteristics of LDIBs as compared to overall neutrophils.

**Fig. 5.**
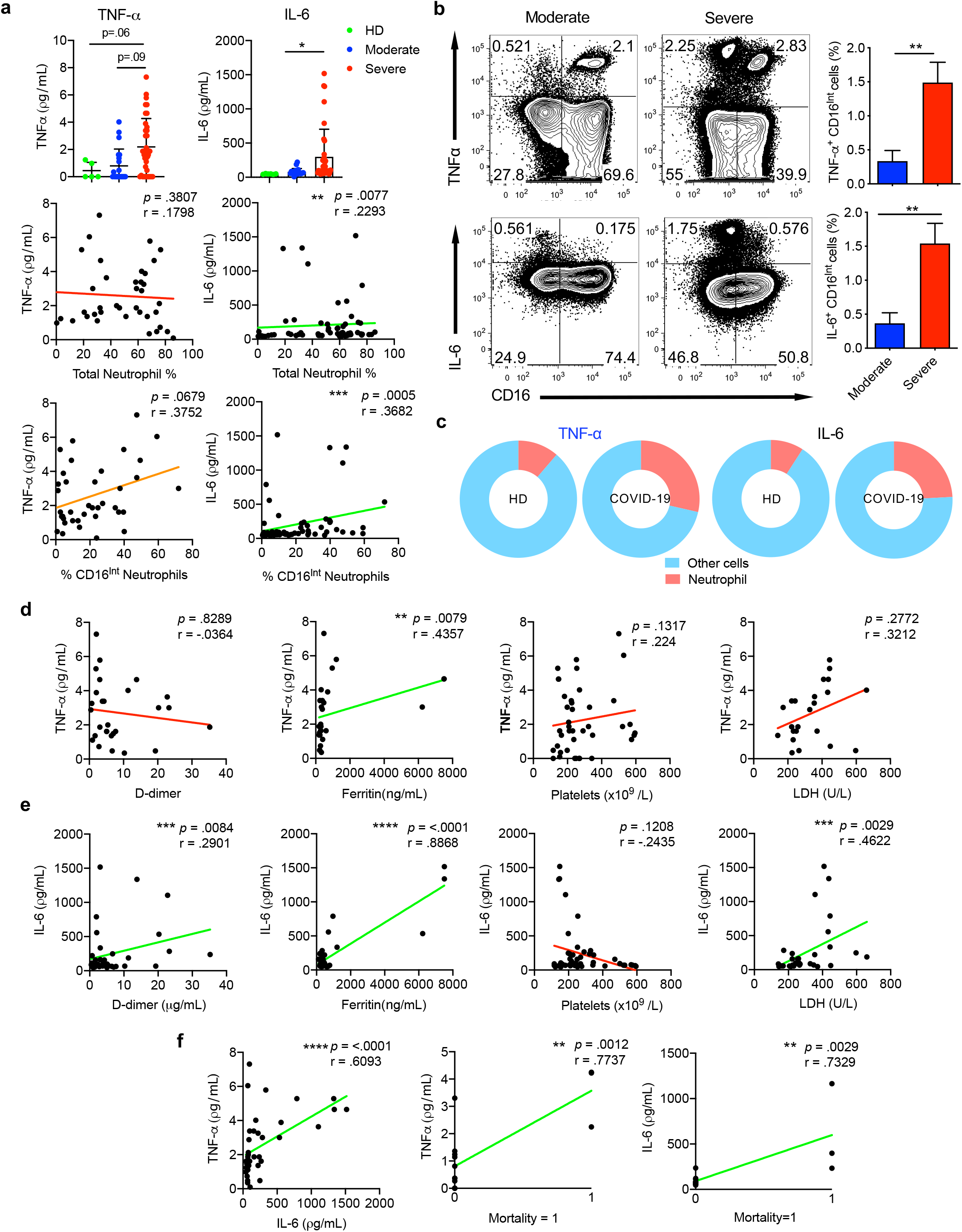
Cytokine production by LDIBs drives clinical features of coagulation. **(A)** An ELISA was used to detect plasma concentrations of IL-6 and TNF-α in each patient sample, HD (n=6), moderate (n=21), severe (n=36) and a mixed linear effect model was used to determine significance between groups. IL-6 and TNF-α levels were then correlated with both the total neutrophil count and the percent of CD16^Int^ neutrophils in the corresponding sample as measured by mass cytometry. Samples that fell below the level of detection of the TNF-α ELISA were excluded from correlation data. (IL-6 n=57, TNF-α n=38) **(B)** Representative plots of TNF-α (top) and IL-6 (bottom) production from LPS stimulated CD16^High^ and CD16^Int^ neutrophils cultured from whole blood samples of moderate (n=4) and severe patients (n=2), with accompanying summarized data. p values were determined using a student’s t-test. **(C)** Pie charts show the relative contribution of neutrophils to the total TNF-α and IL-6 *ex vivo* pool as compared to all other immune cells in healthy donors and COVID-19 patients, indicating an increase in the ratio of TNF-α and IL-6 being made by neutrophils in COVID-19 patients. **(D)** IL-6 plasma concentrations measured in A were also correlated with the clinically measured D-dimer levels from the same day that the sample was acquired (n=38), Ferritin (n=38), Platelets (n=50), and LDH (n=35) **(E))** TNF-α plasma concentrations measured in A were also correlated with the clinically measured values from the same day that the sample was acquired. Samples that fell below the level of detection of the TNF-α ELISA were excluded from correlation data. D-dimer (n=28), Ferritin (n=27), Platelets (n=41), and LDH (n=23) **(F)** Serum concentrations of IL-6 and TNF-α were also correlated with one other (n=38). Patient mortality was correlated with plasma TNF-α and IL-6 concentrations using the mean TNF-α and IL-6 level from a patient’s samples. Patient mortality was indicated in a binary variable where 1 indicated mortality and 0 was used for non-mortality. For all correlation data, a line of best fit is shown to visually examine correlation, with a green line representing a statistically significant correlation, a red line representing a non-significant correlation and an orange line representing a trending correlation that was not significant. Marginal Pearson correlation where used to indicate statistical significance in all correlations, where ns= p ≥.05, * p< 0.05, ** p<0.01 ***, p<0.001, **** p < 0.0001.

We next sought to examine whether neutrophils directly contribute to these systemic cytokine pools. Stimulation of whole blood samples with LPS showed LDIBs in the severe patients were capable of producing significant amounts of TNF-α and IL-6 compared to moderate patients (Figure 5b). In addition, neutrophils from all COVID-19 patients increased their proportion of total cytokine-producing cells compared to those from healthy donors (Figure 5c). Further investigation into the correlation of TNF-α levels with other clinical markers of inflammation demonstrated a significant correlation with ferritin but no correlation with D-dimer, platelets and LDH (Figure 5d). In contrast, IL-6 levels were positively correlated with the levels of D-dimer, ferritin and LDH and negatively trending with platelets (Figure 5e). Furthermore, these two cytokines correlated with each other and both TNF-α and IL-6 significantly correlated with patient mortality (Figure 5f). Overall, these data suggest that neutrophils, particularly the CD16^Int^ LDIB subset, are substantial contributors to the cytokine storm seen in COVID-19 patients. In patients with severe elevations in LDIBs or “LDIB crisis”, the dramatic increase in production of TNF-α and IL-6 likely causes a profound upregulation of tissue factor resulting in thrombus formation and D-dimer elevation.

## Discussion

Our study aimed to investigate the etiology of CAC in an effort to help guide patient management and improve survival outcomes. On average, approximately one third of critically ill COVID-19 patients develop CAC and thromboembolic complications during the course of the disease(*20, 40*). The most common primary outcomes are venous thromboembolism, ischemic stroke, myocardial infarction, and disseminated intravascular coagulation(*20*). In our own patient cohort, 8/13 (61.5%) of COVID-19 patients experienced a thromboembolic complication. Clinically, the majority of severe COVID-19 patients present with grossly elevated D-dimers(*17*). Treating high risk patients with a full dose of systemic anti-coagulation has been shown to be associated with a decreased risk in mortality(*32*). However, systemic anti-coagulation poses potential bleeding risks and is contra-indicated in some patients, especially those with numerous co-morbidities, which make up a significant portion of COVID-19 patients. Additionally, treating the coagulopathy targets the symptoms rather than the cause of the problem.

It has been proposed that the strong inflammatory response to COVID-19 is associated with CAC(*17*). One case study found that IL-6 levels significantly correlated with fibrinogen levels in mechanically ventilated COVID-19 patients(*41*). However, while this suggests that the unchecked inflammatory response could be contributing to CAC, the specific cellular etiology and mechanism have not been directly elucidated. One of the most notable immune disturbances in severe COVID-19 is neutrophilia and increased NLR. Both increased D-dimer and NLR have been associated with poor clinical outcomes(*10, 21*). Therefore, we examined the possibility that the neutrophils are significantly contributing to the coagulopathy and could be used as an adjunct clinical measure to determine thromboembolic complication risk and guide treatment measures.

In agreement with previous reports, we found that severe COVID-19 patients have an increased neutrophil percentage and increased NLR. Here, we further detail the emergence of a novel immature neutrophil population, LDIBs, in the peripheral blood of the severe COVID-19 patients. These cells are identified by their distinct band shaped nucleus in addition to intermediate expression of CD11b and CD16, low expression of CD44 and high expression of CD40 (CD16^Int^CD44^Low^CD11b^Int^). Like low-density neutrophils described in other inflammatory immune conditions, we were able to isolate these cells vial PBMC Ficoll pull down in COVID-19 patients but not in healthy donors(*25*). In accordance with previous reports, these cells readily make NETs which we captured via Wright Giemsa staining. In addition, CD40^+^LDIBs correlate strongly with plasma levels of D-dimer and ferritin in severe COVID-19 patients. Overall, the combination of NET formation and CD40 expression indicates a neutrophil that is capable of promoting coagulation and thrombosis from CD40 mediated platelet activation and NET induced endothelial damage. Additionally, the down regulation of CD44 enables these cells to traffic to the lung where multiple published case studies demonstrate marked neutrophil infiltration into the lung tissue and subsequent damage(*12, 18*). Neutrophil infiltration of the lung is accompanied by lung edema, endothelial injury and epithelial injury, which are hallmark events in the development of ARDS. Hence, the recruitment of LDIBs to the lung in COVID-19 likely plays an important role in the progression of ARDS observed in the most severe patients(*42*) as proposed in our schematic model (Figure 5). Increases in LDIB populations over baseline are also shown to be associated with intubation or patient mortality in our study. Conversely, a decrease in LDIB percentage frequently accompanies a positive clinical prognosis, with extubation or discharge.

Further examination into the functionality of these cells revealed a propensity for spontaneous NET formation and increased secretion of TNF-α and IL-6. Correlating these cells with clinical coagulation factors revealed that LDIBs trended with all COVID-19 patient D-dimer levels and serial analyses of patients’ individual LDIB populations showed apparent associations with D-dimer. LDIB percentage also correlated with systemic IL-6 and TNFα levels as well. It is worth noting that some of these correlation analyses did not reach statistical significances. Many factors could contribute to these results. For example, our patient cohort is relatively small and many parameters such as D-dimer were not frequently measured in the clinical lab work. Nevertheless, our data suggest that LDIBs, at least in part, contribute to CAC through increased secretion of IL-6 and TNF-α particularly during LDIB crisis which results in activation of the extrinsic coagulation cascade causing thrombus formation.

In this study, we used serial patient samples taken during the length of patient hospitalization and grouped these based on the status (moderate or severe) of the patient at that time. In this way we could better capture the dynamic nature of COVID-19 in patients, and better understand how neutrophils and LDIBs change as individual patient’s conditions both improve and deteriorate, and understand how severe versus moderate patients generally differ. In order to then conduct proper statistical analyses, we used linear mixed and marginal Pearson analyses(*43, 44*) to properly account for the use of these serial measurements from patients, as explained in the methods.

Recent publications in the field have called for the use of anti-inflammatory agents in the treatment of COVID-19(*35, 45*). Numerous case reports have shown that COVID-19 patients with a history of inflammatory autoimmune diseases like rheumatoid arthritis or inflammatory bowel disease have a milder course of infection(*46, 47*). However, in the context of the data presented here, the reduced disease severity could be a result of either drug induced neutropenia which is common in autoimmune patients or a result of decreased TNFα/IL-6 levels from monoclonal antibody treatment. There was some hesitation in the field to use immunosuppressive agents like tocilizumab, adalimumab, and etanercept due to concerns about restraining immune function during viral infection(*48*). The challenge remained in correctly identifying the patients who could benefit from immunosuppressive anti-IL-6 and anti-TNF-α therapy versus those in who it may cause delayed viral clearance resulting in worse clinical outcomes. Based on the data we present in this paper, we propose that immunosuppressive agents like tocilizumab and adalimumab, used in conjugation with anti-viral agents, could be beneficial for severe patients in or trending towards LDIB crisis to limit the deleterious effects of these cytokines on inducing coagulation. These patients can be best identified clinically by monitoring the percentage of LDIBs on routine CBCs. Obtaining a serum IL-6 level could further confirm whether a patient is trending towards an LDIB and coagulation crisis. Intervening early before patients hit this crisis could help prevent thromboembolic complications and improve all-cause mortality rates for COVID-19 patients.

## Materials and Methods

### Study Participants and Clinical Data

The Institutional Review Board at University of Louisville approved the present study and written informed consent was obtained from either subjects or their legal authorized representatives (IRB No. 20. 0321). Inclusion criteria were all hospitalized adults (older than 18) at the University of Louisville Health who have positive COVID-19 results and were consented to this study. Exclusion criteria included age younger than 18 and refusal to participate. COVID-19 patients enrolled in this study were diagnosed with a 2019-CoV detection kit using real-time reverse transcriptase-polymerase chain reaction performed at the University of Louisville Hospital Laboratory from nasal pharyngeal swab samples obtained from patients.

The grouping of COVID-19 patients into Moderate Group vs. Severe Group is based on the initial clinical presentation at the time of enrollment. Severe Group participants were COVID-19 confirmed patients who required mechanical ventilation and this group had blood draw daily along with their standard laboratory work. Moderate Group participants were COVID-19 confirmed patients who were hospitalized without mechanical ventilation and had blood draw every two to three days along with their standard laboratory work. All COVID-19 patients were followed by the research team daily and the clinical team was blinded to findings of the research analysis to avoid potential bias.

The demographic characteristics (age, sex, height, weight, Body Mass Index (BMI), clinical data (symptoms, comorbidities, laboratory findings, treatments, complications and outcomes) and results of cardiac examinations including biomarkers, ECG and echocardiography were collected prospectively by two investigators (JH and MW). All data were independently reviewed and entered into the computer database (CW and DT). The clinical outcomes (discharge, mortality and length of stay) were monitored up to May 15, 2020. For hospital laboratory CBC tests, normal values are the following: white blood cell (4.1-10.8 ×10^3^/μL); hemoglobin (13.7-17.5gram/dL); platelet (140-370 ×10^3^/μL). For hospital laboratory inflammatory and coagulation markers, normal values are the following: D-dimer (0.19-0.74 μgFEU/ml); ferritin (7-350 ng/ml); lactate dehydrogenase (LDH) (100-242 Units/Liter).

### Plasma and PBMC Isolation

Whole blood samples were centrifuged at 1600 rpm for 10 minutes. Plasma was aspirated and aliquoted into 1mL Eppendorf tubes and immediately stored at −8°C until future use. The remaining cell layers were diluted with an equal volume of complete RPMI1640. The blood suspension was layered over 5mL of Ficoll-Paque (Cedarlane Labs, Burlington, ON) in a 15mL conical tube. Samples were then centrifuged at 2,000 rpm for 30 minutes at room temperature (RT) without brake. The mononuclear cell layer was then transferred to a new 15mL conical tubes and resuspended in 40mL of RPMI, mixed, and centrifuged at 1,500 RPM for 10 minutes at 4°C. The supernatant was removed and cells were again washed with RMPI1640. The cell pellet was resuspended in 3mL of RPMI1640 and counted for sample processing.

### Whole Blood Analysis

For whole blood analysis, 150uL of whole blood was lysed with 2mL of ACK for 10 minutes. Cells were spun down and washed once with PBS. Cells were then stained with Viability Dye/APC-Cy7, CD45-PeCy7, CD66b-PE, and CD-16 FITC for 30 minutes at 4°C prior to washing and analysis of a BD FACS Canto.

### *Ex vivo* neutrophil stimulation

Whole blood (150uL) was lysed with ACK buffer. One-million cells were seeded in a 24-well plate and cultured with Brefeldin A solution for 20 minutes at 37°C. Cells were then stimulated with 250ng/mL of LPS for 10 hours at 37°C. Following stimulation, cells were collected and washed with PBS prior to cell surface staining with Viability Dye-APC-Cy7, CD45-PE-Cy7, CD66b-PE, CD16-APC for 30 minutes at 4°C. Cells were washed again with PBS before fixation (Biolegend Intracellular Fixation Buffer) for 30 minutes at RT. Cells were then washed twice with permeabilization buffer (Biolegend Per Wash Buffer). Cells were incubated with TNFα-PerCP-Cy5.5 and IL-6-FITC overnight prior to washing and analysis on BD FACS Canto.

### Wright Giemsa Stain

Half-million PBMCs were stained with Viability Dye-APC-Cy7, CD45-PerCP-Cy5.5, CD66b-PE, CD16-APC for 30 minutes at 4°C prior to washing with AutoMACs running buffer. Cells were then sorted based on CD16 expression using a BD FACS Aria III. Following collection, cells were spun down at 1600 RMP for 8 minutes. Cells were resuspended in 200uL and spun onto a microscope slide (800 rpm for 5 minutes) using a Shandon CytoSpin3 (Thermo Fisher). Slides were then air dried for 10 minutes prior to staining. For the Wright Giemsa Stain (Shandon Wright Giemsa Stain Kit, Thermo Fisher), slides were dipped in Wright-Giemsa Stain Solution for 1 minute and 20 seconds. After blotting off excess stain, slides were dipped in Wright Giemsa Buffer for 1 minute and 20 seconds. Slides were blotted to remove excess buffer. Slides were then dipped into the Wright-Giemsa Rinse Solution for 10 seconds using quick dips. The back of the slides were wiped and set to dry in a vertical position for 10 minutes prior to analysis on an Aperio Scan Scope.

### CyTOF Mass Cytometry Sample Preparation

Mass cytometry antibodies (Supplemental Table 1) were either purchased pre-conjugated (Fluidigm) or were conjugated in house using MaxPar X8 Polymer Kits or MCP9 Polymer Kits (Fluidigm) according to the manufacturer’s instruction. PBMCs were isolated as described above. The starting cell number was 1.0×10^6^ cells per patient. The samples were stained for viability with 5uM cisplatin (Fluidigm) in serum free RPMI1640 for 5 minutes at RT. The cells were washed with FBS (10%) containing RPMI1640 for 5 minutes at 300xg. Cells were stained with the complete antibody panel for 30 minutes at RT. Cells were then washed and fixed in 1.6% formaldehyde for 10 minutes at RT. They were washed and then incubated overnight in 125nM of Intercalator-Ir (Fluidigm) at 4°C.

### CyTOF Data Acquisition

Prior to acquisition, samples were washed twice with Cell Staining Buffer (Fluidigm) and kept on ice until acquisition. Cells were then resuspended at a concentration of 1 million cells/mL in Cell Acquisition Solution containing a 1/9 dilution of EQ 4 Element Beads (Fluidigm). The samples were acquired on a Helios (Fluidigm) at an event rate of <500 events/second. After acquisition, the data were normalized using bead-based normalization in the CyTOF software. The data were gated to exclude residual normalization beads, debris, dead cells and doublets, leaving DNA^+^CD45^+^Cisplatin^low^ events for subsequent clustering and high dimensional analyses.

### CyTOF Data Analysis

CyTOF data was analyzed using a combination of the Cytobank software package(*49*) and the CyTOF workflow(*50*), which consists of suite of packages(*51*) (*52-55*) available in R (r-project.org). For analysis conducted within the CyTOF workflow, FlowJo Workspace files were imported and parsed using functions within flowWorkspace(*52*) and CytoML(*53*). An arcsinh transformation (cofactor=5) was applied to the data using the dataPrep function within CATALYST(*54*) and stored as a singlecellexperiment object(*55*). Cell population clustering and visualization was conducted using FlowSOM(*56*) and ConsensusClusterPlus(*51*) within the CyTOF workflow and using the viSNE application within Cytobank. Depending on the analysis, clustering was either performed using data across all donors at the first blood draw (Healthy Donors, n=5; Moderate, n=6; Severe, n=7), or using data from selected patients across multiple time points. Additionally, clustering was performed either using all live CD45^+^ cells or after gating on CD66b^+^ neutrophils.

### TNF-α and IL-6 quantification

Plasma concentrations of TNFα and IL-6 were measured using enzyme-linked immunosorbent assay (ELISA) kits (BioLegend, San Diego, CA). The operating procedure provided by the manufacturer was followed. One-hundred μL of plasma was used for each sample. The optical density (OD) at 450 nm was measured using a Synergy™ HT microplate reader (BioTek, Winooski, VT). Concentrations of TNF-α and IL-6 were determined using the standard curves. A few OD readings fell outside of the range of the standard curve, in which case a line of best fit was used to extrapolate the data.

### Phagocytosis Assay

Cells were acquired from whole blood following ACK lysis. One-million cells were washed with HEPES diluted 50x in RPMI1640, and then incubated in 100μL of this solution for 1 hour at 37°C for activation. The pHrodo™ Green S. *aureus* BioParticles™ Phagocytosis Kit for Flow Cytometry was used, where 100μL of the reconstituted particles were added to the activated whole blood, and incubated for 1 hour at 37°C. Samples were lightly mixed every 20 minutes. The reaction was stopped with 1mL of cold PBS, and surface staining for viability, CD45, CD66b and CD16 (BioLegend, San Diego, CA) was performed. Samples were analyzed using a BD FACSCanto (BD Biosciences, Oxford, UK), and cells that fluoresced in the FITC channel were determined to be phagocytic.

### Statistical analysis

First descriptive statistics such as mean and standard deviation and graphics were presented for each variable, stratified by study groups. Since we have varied number of observations for each patient, we applied linear mixed effect models along with the Wald test statistics to compare the group differences(*44*), where group was considered as fixed effects, and patients were considered random effects. To examine association between two variables, we estimated the marginal Pearson correlation coefficient and tested its significance.(*43*) The marginal Pearson correlation coefficient captures the association between two variables at the population level. The analyses were carried out in the Statistical software R (https://www.r-project.org/) and Prism version 10. A statistical test was claimed significant if p< 0.05.

## Data Availability

Yes all raw data are available upon request

## Acknowledgements

The authors would like to thank Dr. Ozan Akca, Dr. Alexander F. Bautista, and Mr. Zachary Martin (medical student at University of Louisville) for their help on patient recruitment and study implementation. We would also like to thank Drs. Kelly M. McMasters and Michael Kommor for his feedback on this project. Finally, the authors would like to acknowledge and thank the healthy donors and the COVID-19 patients and their families for participating in this study. For those who lost a family member or loved one, we are so very truly sorry for your loss. The Bill and Lindy Street Gift Fund provided support for the CyTOF acquisition and operation.

## Author Contributions

SM, AG, and XH lead data collection and analysis. CW and DT performed data analysis for clustering algorithms. JY, JH, CD, HZ, and LC contributed to experimental design. EC, RC, SC, JC, and JH consented and recruited patients. JH and MW performed patient chart review. MK performed and supervised all biostatistical analysis. SM, AG, and JY wrote the manuscript. CW, LC, MK, and JH edited and revised the manuscript. JH and JY designed the study, implemented the experiments, and supervised the data analysis.

## Inventory for Supplemental materials

1. Supplemental Table 1

2. Five Supplemental Figures

**Supplemental Table 1.** Mass cytometry antibody panel

| Antigen | Symbol and Mass | Antibody clone | Source |
| --- | --- | --- | --- |
| CD45 | 89Y | HI30 | Fluidigm |
| CD8 | 106Cd | RPA-T8 | Biolegend-Custom |
| CD14 | 110Cd | M5E2 | Biolegend- Custom |
| CD4 | 111Cd | RPA-T4 | Biolegend-Custom |
| CD11b | 112Cd | IRCF44 | Biolegend-Custom |
| CD3 | 113Cd | UCHT1 | Biolegend-Custom |
| CD20 | 114Cd | 2H7 | Biolegend-Custom |
| CD19 | 116Cd | HIB19 | Biolegend-Custom |
| CD196 | 141Pr | G034E4 | Fluidigm |
| CD40 | 142Nd | 5C3 | Fluidigm |
| CD123 | 143Nd | 6H6 | Fluidigm |
| CD69 | 144Nd | FN50 | Fluidigm |
| CD163 | 145Nd | GHI/61 | Fluidigm |
| IgD | 146Nd | IA6-2 | Fluidigm |
| CD11c | 147Sm | Bu15 | Fluidigm |
| CD66b | 148Nd | G10F5 | Biolegend-Custom |
| CD45RO | 149Sm | UCHL1 | Fluidigm |
| LAG-3 | 150Nd | 11C3C65 | Fluidigm |
| LAMP1 | 151Eu | H4A3 | Fluidigm |
| CD21 | 152Sm | BL13 | Fluidigm |
| $\gamma\delta$TCR | 153Eu | B1 | Biolegend-Custom |
| TIM-3 | 154Sm | F38-2E2 | Fluidigm |
| CD56 | 155Gd | HCD56 | Fluidigm |
| CD86 | 156Gd | IT2.2 | Fluidigm |
| TLR4 | 158Gd | HTA125 | Fluidigm |
| CD197 | 159Tb | G043H7 | Fluidigm |
| CD28 | 160Gd | CD28.2 | Fluidigm |
| CD80 | 161Dy | 2D10.4 | Fluidigm |
| CD79b | 162Dy | CB3-1 | Fluidigm |
| CXCR3 | 163Dy | G025H7 | Fluidigm |
| CXCR5 | 164Dy | RF8B2 | Fluidigm |
| CD45RA | 165Ho | HI100 | Biolegend-Custom |
| CD44 | 166Er | BJ18 | Fluidigm |
| CD27 | 167Er | L128 | Fluidigm |
| CD40L | 168Er | 24-31 | Fluidigm |
| CD25 | 169Tm | 2A3 | Fluidigm |
| CTLA-4 | 170Er | 14D3 | Fluidigm |
| CD68 | 171Yb | Y1/82A | Fluidigm |
| CD38 | 172Yb | HIT2 | Fluidigm |
| HLA-Dr | 173Yb | L243 | Fluidigm |
| CD279 | 174Yb | EH12.2H7 | Fluidigm |
| CD274 | 175Lu | 29E.2A3 | Fluidigm |
| CD127 | 176Yb | A019D5 | Fluidigm |
| CD16 | 209Bi | 3G8 | Fluidigm |

**Figure S1.**
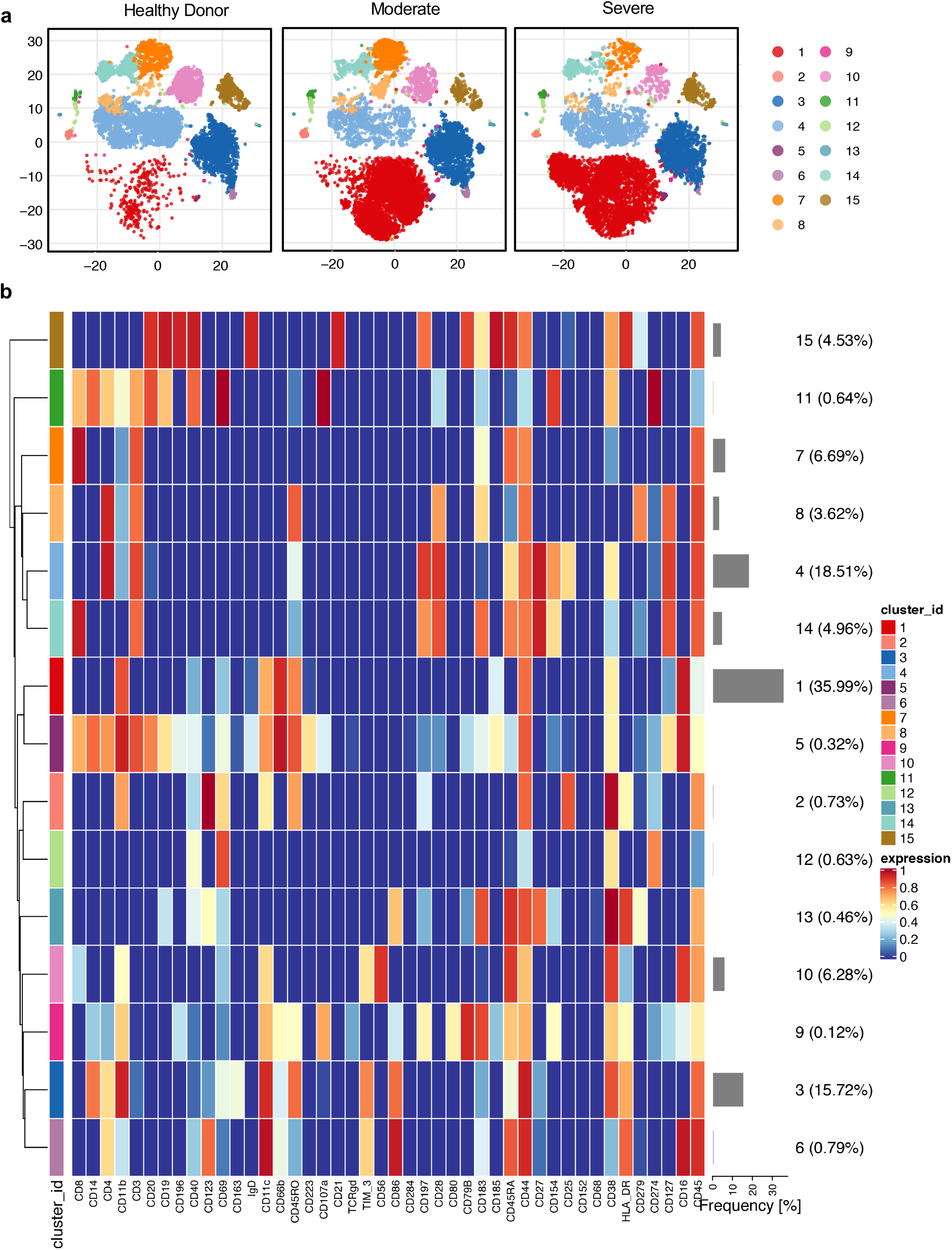
Cluster analysis of CD45^+^ PBMCs in healthy donors, moderate and severe COVID-19 patients. **(A)** Representative cluster maps for moderate and severe COVID-19 patients as compared to healthy donors. The data was generated from CyTOF based analysis of CD45^+^ PBMCs isolated from peripheral blood. **(B)** Heatmap of differential expression pattern of lineage and surface markers in PBMCs of moderate and severe COVID-19 patients as compared to healthy donors. The color key identifies the cluster populations shown above. Here, 5 HDs, 5 moderate COVID-19 patients and 6 severe COVID-19 patient samples from the first day of study enrollment were used to generate the plots.

**Figure S2.**
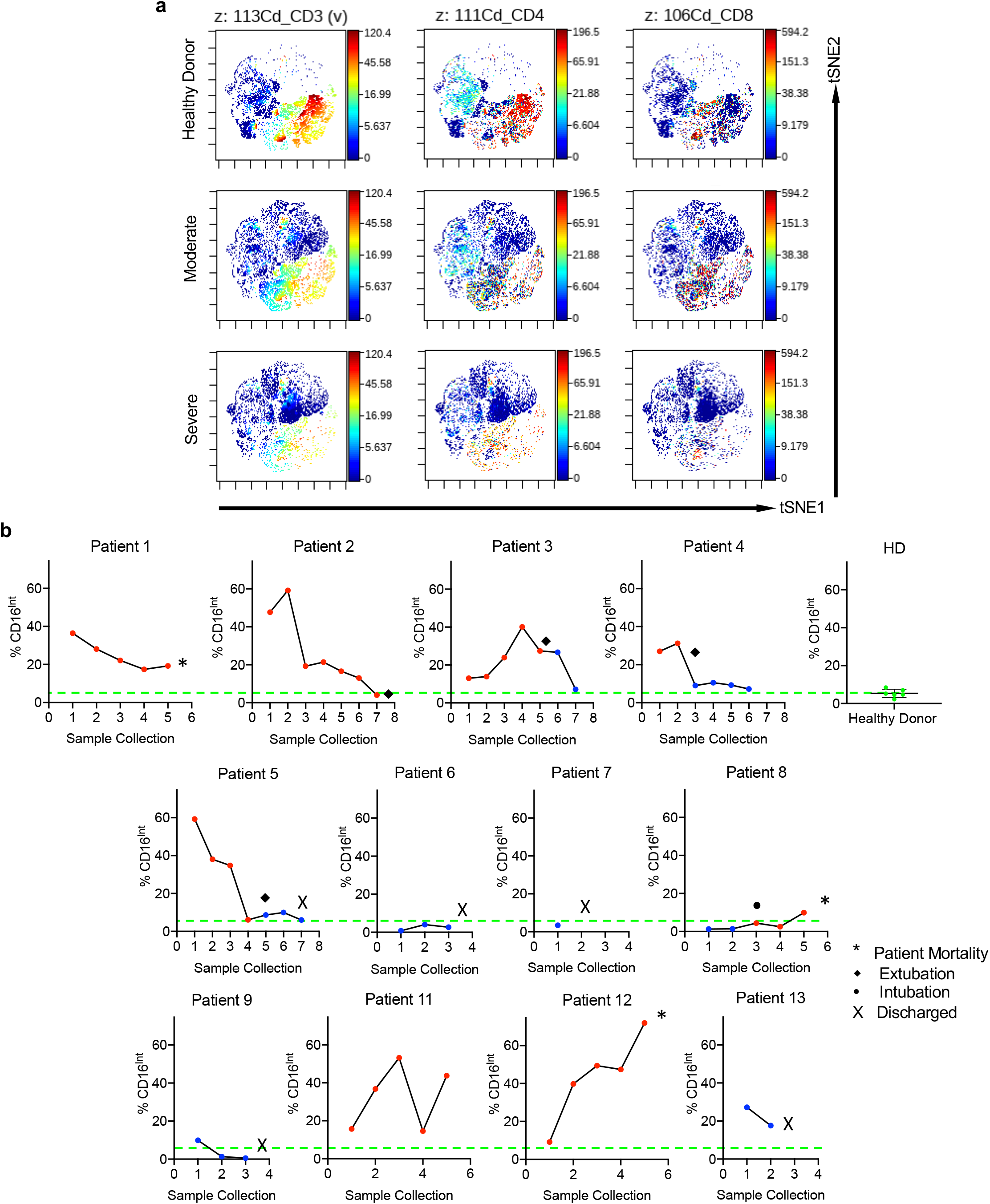
Longitudinal immune profiling of moderate and severe COVID-19 patients. **(A)** Representative viSNE plots generated using CytoBank showing decreased CD3 (left), CD4 (middle), and CD8 (right) expression in moderate and severe COVID-19 patients as compared to healthy donors in the CD45^+^ compartment of PBMCs. Here, 5 HDs, 5 moderate COVID-19 patients and 6 severe COVID-19 patient samples from the first day of study enrollment were used to generate the plots. **(B)**. Serial blood draws from our patient cohort enables us to track the CD16^Int^ LDIB population percentage in Ficoll isolated PBMCs over the course of patient hospitalization and correlate it with patient severity and in some cases, clinical outcomes. The first time point indicates enrollment into our study. For the severe patient cohort, samples were collected and analyzed everyday whereas in the moderate cohort, on average, samples were obtained every third day. A red dot indicates that a patient is classified as severe whereas a blue dot signifies a patient is considered moderate. The green line represents the average level of CD16^Int^ neutrophils in healthy patients for a reference of a “normal” level.

**Figure S3.**
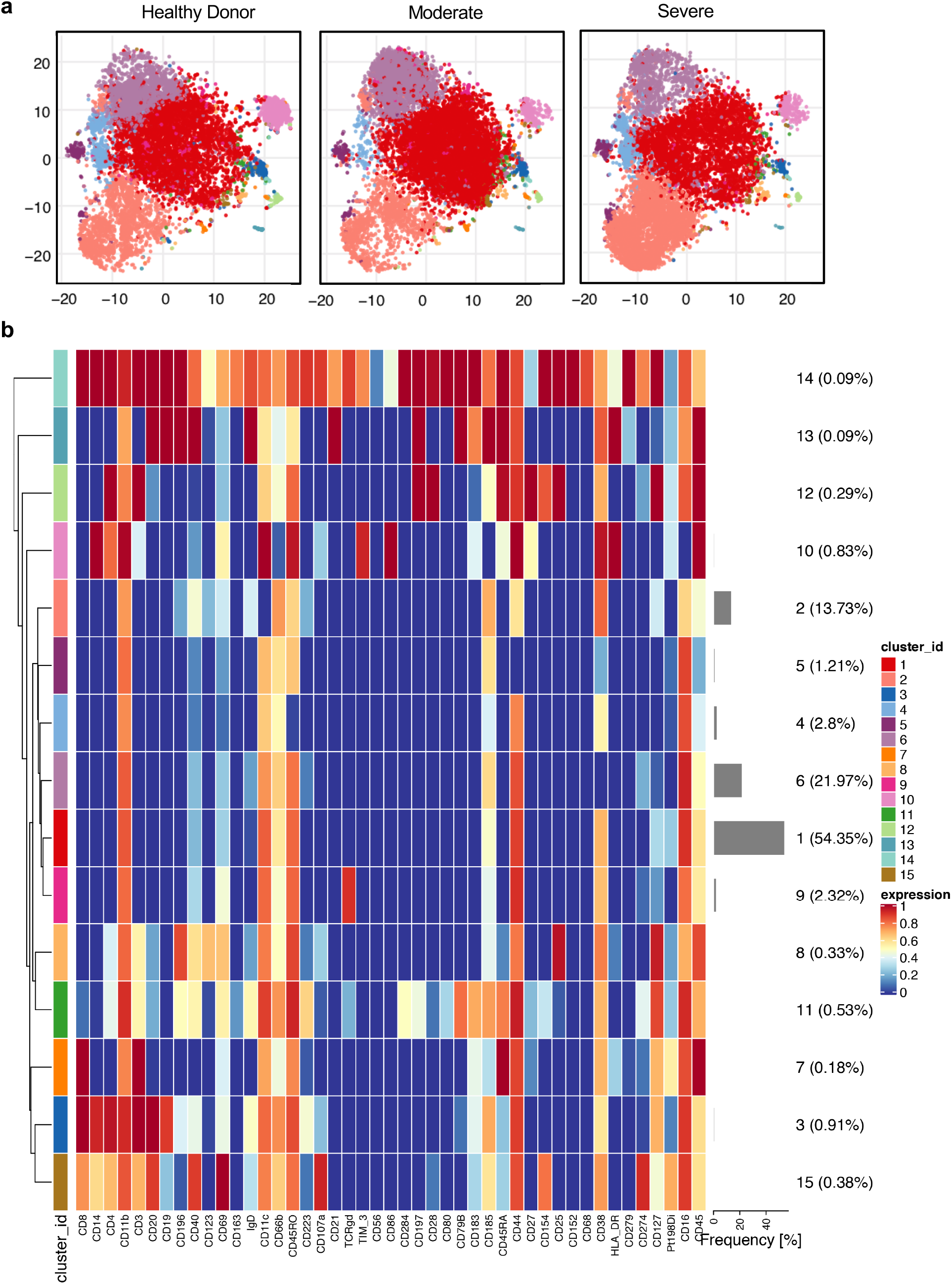
Surface marker expression profiling of neutrophils in moderate and severe COVID-19 patients. **(A)** Representative cluster maps of neutrophil subsets in moderate and severe COVID-19 patients as compared to healthy donors. Here, data from 5 HDs, 5 moderate COVID-19 patients and 6 severe COVID-19 patient samples from the first day of study enrollment were used to generate the plots. **(B)** Heatmap showing differential surface marker expression of the overall CD66b^+^ neutrophil populations in moderate and severe COVID-19 patients as compared to healthy donors.

**Figure S4.**
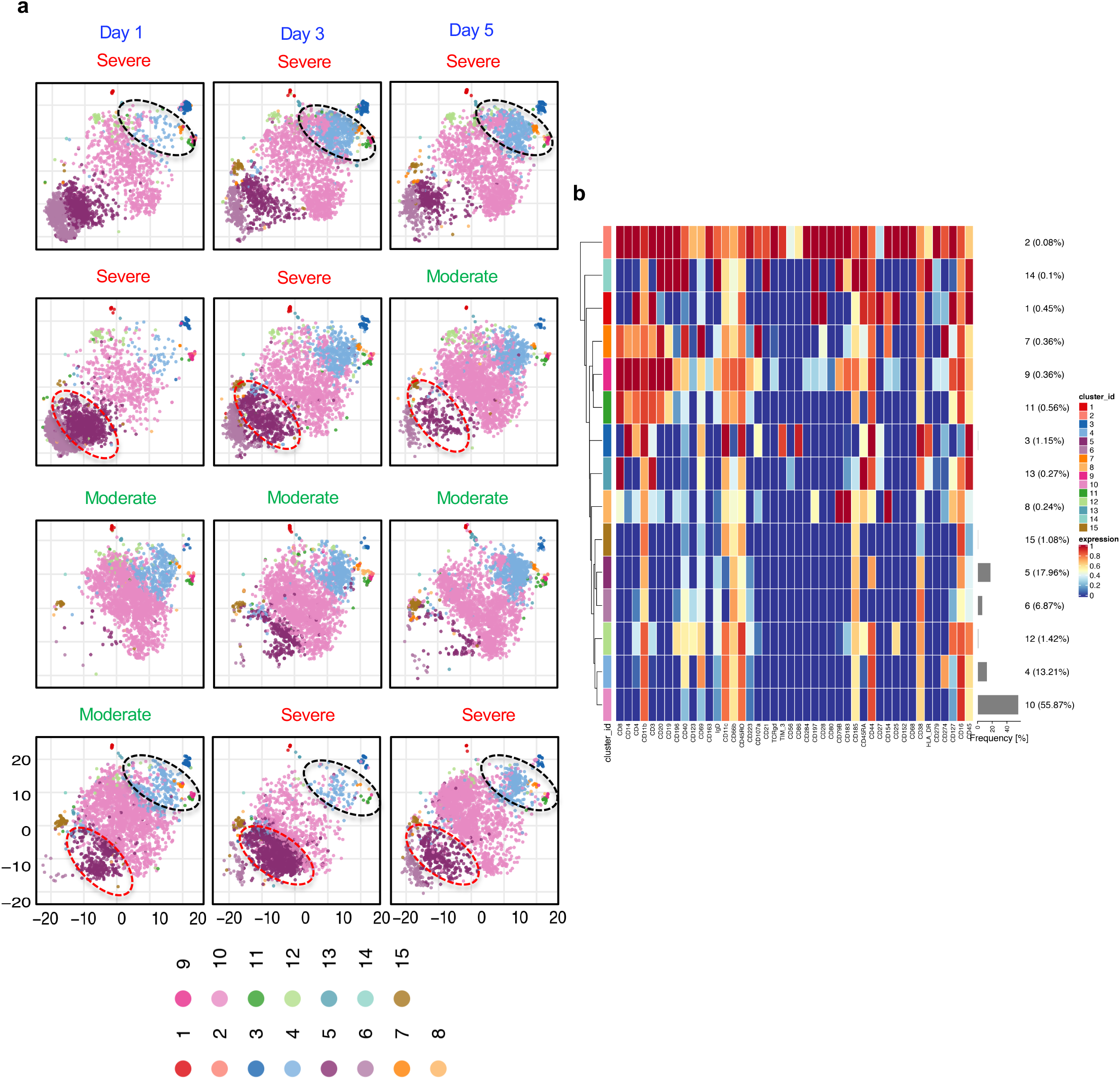
Differential expression of neutrophil clusters in patients over their clinical course of disease. **(A)** viSNE plots representing the total CD66b^+^ neutrophil pool in 4 patients who experienced different clinical courses from days 1, 3 and 5 of study enrollment. Data represents a patient who was classified as severe on days 1, 3 and 5 (top), a patient whose condition improved, and was transitioned to a moderate patient by day 5 (2^nd^ from top), a patient who remained in the moderate group for the entirely of the study (2^nd^ from bottom), and one patient who progressed from the moderate to severe group (bottom). The dynamic nature of CD66b^+^ neutrophil populations over the course of disease are highlighted by the black and red circles, where cluster surface marker phenotypes are indicated in S4b. **(B)** Heatmap showing differential surface marker expression on the CD66b^+^ neutrophil pool, which indicates specific subsets of neutrophil populations within the neutrophil compartment.

**Figure S5.**
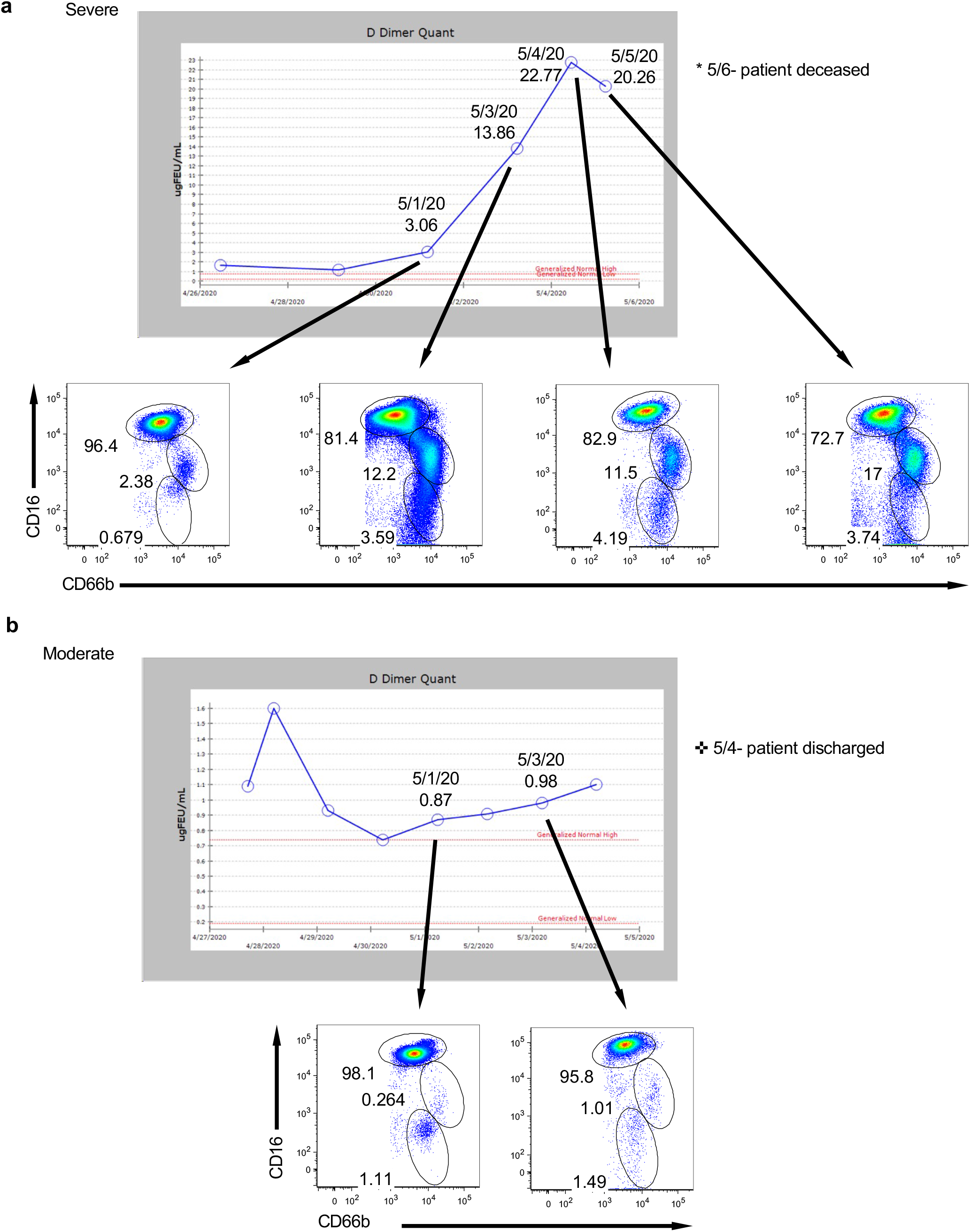
Trending LDIB population with clinical D-dimer levels. Sequential whole blood analysis of the CD16^Int^ LDIB population (middle circle) for severe (**A**) and moderate (**B**) COVID-19 patients overlaid with clinical D-dimer quants from the corresponding days.

